# Vascular Inflammation in Lungs of Patients with Fatal Coronavirus Disease 2019 (COVID-19) Infection: Possible role for the NLRP3 inflammasome

**DOI:** 10.1101/2021.03.19.21253815

**Authors:** Oindrila Paul, Jian Qin Tao, Leslie Litzky, Michael Feldman, Kathleen Montone, Chamith Rajapakse, Christian Bermudez, Shampa Chatterjee

**Affiliations:** Institute for Environmental Medicine and Department of Physiology, University of Pennsylvania School of Medicine, Philadelphia PA 19104; Department of Pathology, University of Pennsylvania School of Medicine, Philadelphia PA 19104; Department of Radiology, University of Pennsylvania School of Medicine, Philadelphia PA 19104; Department of Surgery, University of Pennsylvania School of Medicine, Philadelphia PA 19104

## Abstract

Hyperinflammation is a key event that occurs with SARS-CoV-2 infection. In the lung, hyperinflammation leads to structural damage to tissue. To date, numerous lung histological studies have shown extensive alveolar damage, but there is scarce documentation of vascular inflammation in postmortem lung tissue. Here we document histopathological features and monitor the NLRP3 inflammasome in fatal cases of disease caused by SARS Cov2 (COVID-19). We posit that inflammasome formation along the vessel wall is a characteristic of lung inflammation that accompanies COVID-19 and that it is a probable candidate that drives amplification of inflammation post infection.

## INTRODUCTION

It has been more than a year since the pandemic caused by the novel SARS-CoV-2 corona virus (Severe Acute Respiratory Syndrome Coronavirus), also known as COVID-19 has affected large populations globally (13, 39). The virus disproportionately affects the respiratory system and a major cause of fatality is the acute respiratory distress syndrome (ARDS) that accompanies the infection (10, 19). Autopsy-based lung histological studies have been an invaluable tool in understanding the pathobiology of COVID-19; indeed these have shown indications of inflammation, edema, coagulopathy and fibrosis (6, 10, 14, 36, 40).

COVID-19 manifests itself under a wide spectrum of symptoms, but it can broadly be classified as an inflammatory disease where excessive inflammation is the main driver of poor clinical outcome (8, 15). In this direction, the vascular endothelium, a dynamically adaptable interface that is actively involved in recruitment of inflammatory cells, possibly plays a crucial role in regulation, progression, and amplification of inflammation. While post mortem findings have shown alveolar damage, early or intermediate proliferative phase, and presence of thrombi and signs of inflammation in the lungs (6, 10, 40), histopathology in the context of vascular inflammation and altered vascular structures has been somewhat scarce (1, 36).

Inflammatory processes involve the participation of inflammasomes that are multimeric platforms assembled in response to pathogenic stimuli. Dysregulated inflammasome signaling has been well established as a pivotal event in hyper-inflammatory syndromes (25, 33, 37). Among the inflammasomes, the NLRP3 inflammasome comprising of the NLRP3 subunit, ASC and caspase-1, is well established to be activated in response to microbial infection (2, 29) and to drive cell death (3, 31). It is also involved in COVID-19, as evidenced by the detection of inflammasome subunits and products in the sera and lung tissue of COVID-19 patients (27, 34). However, there are no reports of the presence of the inflammasome in the pulmonary vasculature with COVID-19 infection. As the vasculature seems to be crucial in inflammation accompanying COVID-19, the status of NLRP3 along the vascular wall needs to be documented.

We posit that inflammasome formation is characteristic of pulmonary vascular inflammation that accompanies COVID-19. The purpose of this study is to contextualize vascular features in lung tissue in fatal cases of COVID-19 and ascertain NLRP3 expression along the vascular wall. Here we document the major histological findings of 8 postmortem examinations done on patients with clinically confirmed COVID-19. This study contributes to the growing data on this topic (4, 6, 10, 16, 23, 28).

## MATERIALS AND METHODS

We analyzed lung tissue samples of 8 patients that died of COVID-19 in 2020. Written informed consent was obtained for postmortem examination from the next of kin of these patients. All patients had SARS-CoV-2 infection confirmed by real time PCR analysis at the time of hospital admission. Autopsies were done by trained personnel using personal protective equipment in accordance with the recommendations of the University Of Pennsylvania School Of Medicine.

Tissue blocks taken from the most representative areas of the lung (as identified by macroscopic examination) were fixed in formalin. Paraffin embedded sections of 3 to 5 μm thickness were stained with hematoxylin and eosin (H & E). Images were captured on the Aperio Pathology System and visualized by ImageScope (Leica Biosystems, Buffalo Grove, IL). High and low powered fields were selected for evaluation. Inflammation and inflammation induced cell death (pyroptosis) were characterized by immunostaining for NLRP3 inflammasome and caspase respectively. Sections were deparaffinized; after antibody retrieval, were stained using anti-human NLRP3 monoclonal antibody at 1:200 or anti-human caspase antibody at 1:100 (both from R%D Systems, Minneapolis, MN). Secondary antibody used was conjugated to Alexa 488 at 1:200 (Life Technologies, Eugene, OR). Appropriate IgG controls were used. Vectashield antifade mounting medium used was from Vector Labs (Burlingame, CA). Images were acquired by epifluorescence microscopy using a Nikon TMD epifluorescence microscope, equipped a Hamamatsu ORCA-100 digital camera, and MetaMorph imaging software (Universal Imaging, West Chester, PA, USA). Fluorescence images were acquired at excitation= 488 nm; all images were acquired with the same exposure and acquisition settings as reported previously (5, 7, 32).

## RESULTS

Patient demographics and clinical information are summarized in Table 1 and histological findings in Table 2. Patients were 4 men and 4 women, with a mean age of 71.8 years (SD 13.9). All sections from all patients showed diffuse alveolar damage and intraalveolar fibrin deposition. All sections also stained positively for the NLRP3 inflammasome associated markers that were assessed.

**Table 1:** Patient characteristics, comorbidities, select immunostaining findings on a score of 0 to 3: 0, absent; 1, mild; 2, moderate; 3, severe.

| Patient | Gender | Age | Known Medical History | Substance Abuse<br>(Smoking/Alcohol) | Thrombi/microthrombi | Presence of<br>Inflammation<br>Marker<br>(NLRP3) | Pyroptosis |
| --- | --- | --- | --- | --- | --- | --- | --- |
| 1. | Female | 60s | Asthma and<br>Stroke | Non-smoking | 2 | 2 | 3 |
| 2. | Female | 60s | Breast cancer<br>and therapy<br>related Acute<br>Leukemia | Smoking | 3 | 2 | 3 |
| 3. | Female | 70s | COPD | Smoking and<br>Alcohol | 3 | 2 | 3 |
| 4. | Female | >85 | COPD, Coronary<br>Artery Disease<br>and Sjogrens<br>disease | Not known | 3 | 2 | 3 |
| 5. | Male | 50s | Myeloproliferative<br>disorder and<br>Pulmonary/portal<br>Hypertension | Not known | 3 | 2 | 3 |
| 6. | Male | 70s | Dementia,<br>Diabetes and<br>Hypertension | Not known | 3 | 3 | 3 |
| 7. | Male | 70s | Pulmonary<br>Embolism and<br>Deep Vein<br>Thrombosis and<br>Hypertension | Not known | 3 | 3 | 3 |
| 8. | Male | 80s | Cerebral<br>Vascular Disease | Not known | 3 | 2 | 3 |

**Table 2.** Pulmonary pathological features in COVID-19 autopsy cases on a score of 0 to 4: 0, absent; 1, mild 2, moderate; 3, high; 4, severe.

| Patient No. | Hyaline Membranes | Interstitial Fibrosis | Atypical pneumocytes | Pulmonary hemorrhage | Morphological aspects |
| --- | --- | --- | --- | --- | --- |
| 1. | 4 | 4 | 4 | 3 | Proliferative phase of diffuse alveolar damage, thrombi/microthrombi. |
| 2. | 3 | 3 | 4 | 3 | Emphysematous change, microthrombi, alveolar septal thickening, thrombi/microthrombi. |
| 3. | 4 | 4 | 4 | 3 | Pulmonary edema, alveolar septal thickening |
| 4. | 4 | 4 | 4 | 4 | Proliferative phase of diffuse alveolar damage, pulmonary hemorrhage, thrombi/microthrombi. |
| 5. | 4 | 4 | 4 | 3 | Diffuse alveolar damage, Advanced proliferative phase, thrombi/microthrombi. |
| 6. | 4 | 4 | 4 | 4 | Advanced proliferative phase, pulmonary hemorrhage, thrombi/microthrombi. |
| 7. | 4 | 4 | 4 | 4 | Exudative phase diffuse alveolar damage, hemorrhage, thrombi/microthrombi. |
| 8. | 4 | 4 | 4 | 4 | Advanced proliferative phase, hemorrhage, thrombi/microthrombi. |

Upon light microscopic examination, the lungs of all patients showed extensive alteration of lung microstructure, were patchy and congested (Figure 1A); closer inspection revealed fibrin exudation into alveolar space, thrombi and fibroblastic proliferation, hyaline membrane, fibrin deposition, and early and advanced proliferative phase of diffuse alveolar damage (Figure 1B). Thrombi and microthrombi were identified in 7 of the 8 patients (Figure 2A). Vascular thrombi in small vessels were associated with areas of diffuse alveolar damage. Microthrombi were observed in alveolar septa. Thrombi and microthrombi were widespread in all the samples examined and found in >75% of the fields imaged. Most vessels were obliterated as shown in Figure 2 B. Histological findings are detailed in the legends and in Table 2.

**Figure 1.**
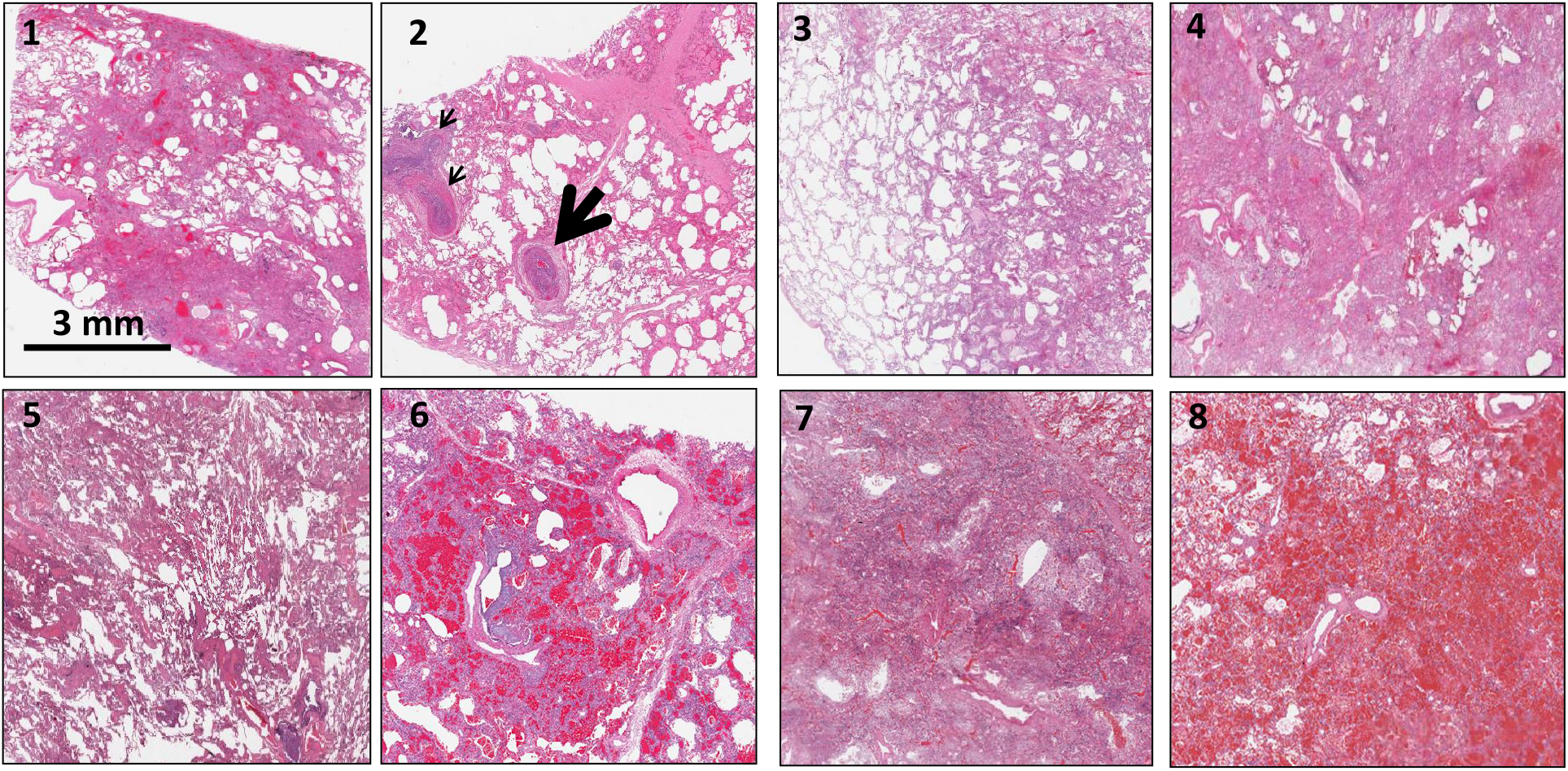
Hematoxylin and Eosin stained sections staining from representative regions of the lung parenchyma of post-mortem lung tissue of 8 patients. A. All patients show extensive alteration of lung microstructure in the form of alveolar damage, fibrin exudation into alveolar space, thrombi and fibroblastic proliferation. The septa are thickened and there is presence of hyaline membranes and dense infiltrates. Scale bar is 3 mm. 1: Alveolar damage with collagen deposition and exudative pattern of damage 2. Large thrombi and smaller caliber arteries showing fibrin thrombi (arrows) 3. Alveolar damage pattern arising from fibroblastic proliferations 4 and 5. Exudate in the entire lung 6. Necrosis with blood and exudate in the lung parenchyma 7. Hemorrhagic infarction of lung tissue adjacent to a pulmonary artery with thrombotic material 8. Pulmonary hemorrhage with blood and fibrin exudation into the parenchyma

**Figure 1 B.**
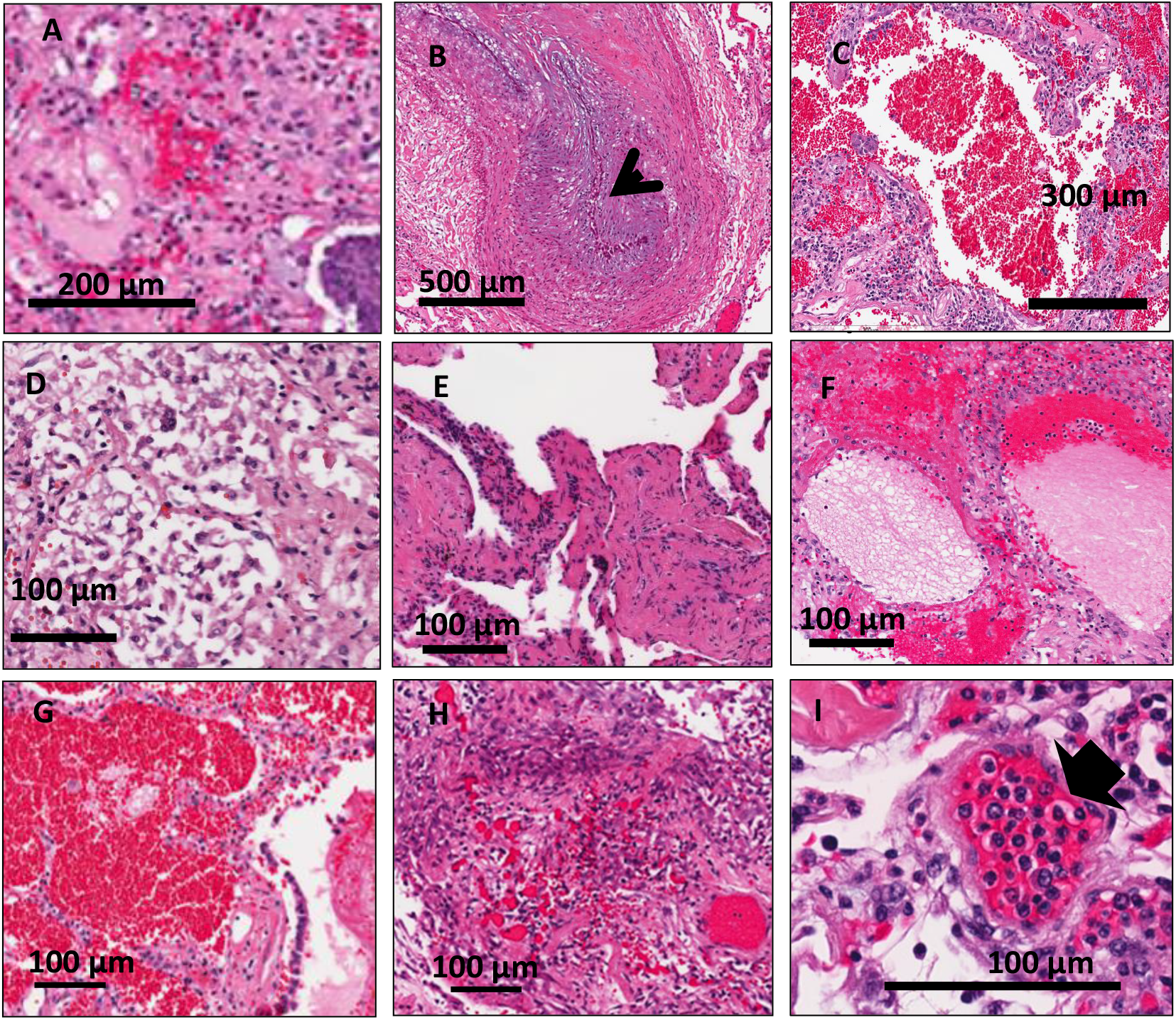
H and E staining at higher magnification: All patients had extensive diffuse alveolar damage, microthrombi and edema in regions of the lung. A. Fibroblastic proliferation B. Plugged airway due to remodeling C. Coagulation necrosis with blood in the lung tissue D. Proliferative phase of diffuse alveolar damage E. Patchy distribution of damage F. Proteinaceous exudates in alveolar spaces G. Blood and fibrin exudation into parenchyma H: Fibroblastic proliferation I: Endotheliitis of small vessel <100 μm with infiltration of the vessel wall by lymphocytes (arrow shows infiltrated cells)

**Figure 2 A.**
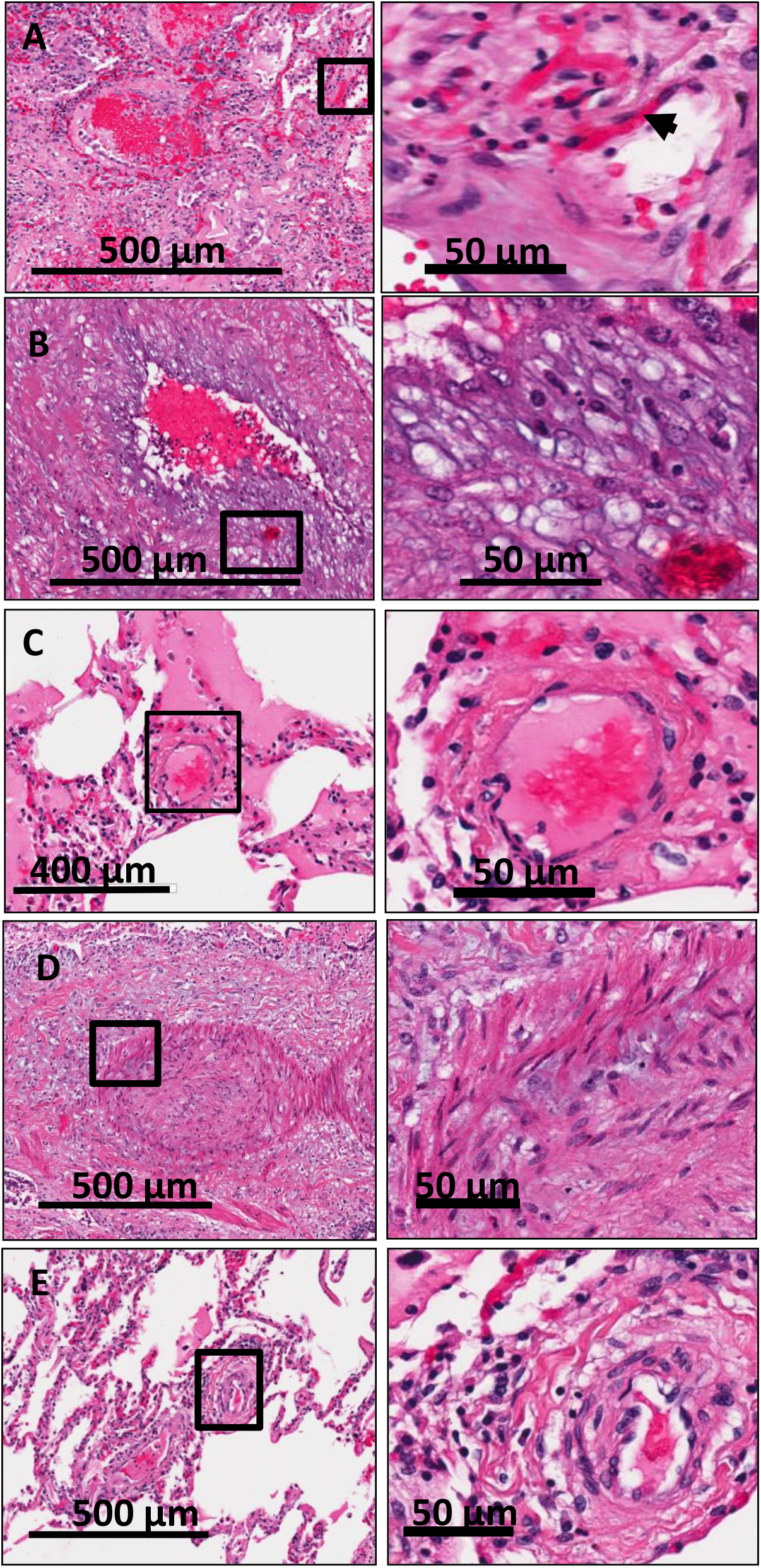
Thrombi and microthrombi were identified in 7 of the 8 patients. Box is magnified in the right panel. Arrow shows microthombi on alveolar septa.

**Figure 2 B.**
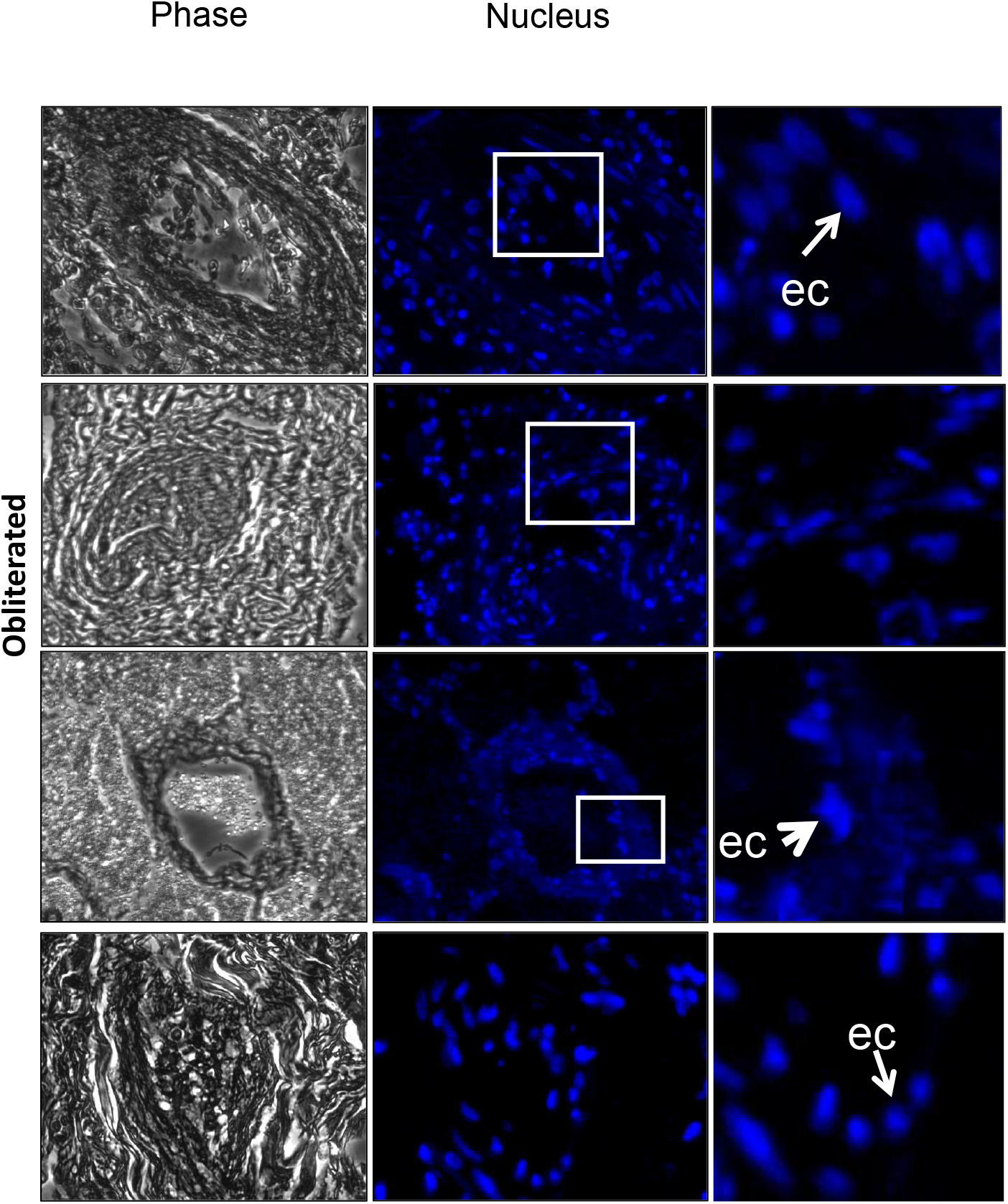
Representative Images of Obliterated Lung Vessels (Phase) indicate that these are either blocked (middle panel) or there is presence of proteinaceous exudates. Nuclear stain (Blue: DAPI) shows the presence of cells inside the vessels. Inset is magnified to show cells possibly endothelial cells (ec) as they are along the wall. But these are few and far between.

We next assessed the expression of the NLRP3 subunit and caspase-1 in all samples. Intense expression of the NLRP3 and caspase-1, as observed from the green fluorescent signal is shown in Figure 3. Fluorescence around the vessel walls implied NLRP3 expression along the endothelial layer (Figure 3A). In contrast, the effector enzyme, caspase-1 was widely distributed throughout the lungs and was not limited to the vascular structures (Figure 3B).

**Figure 3.**
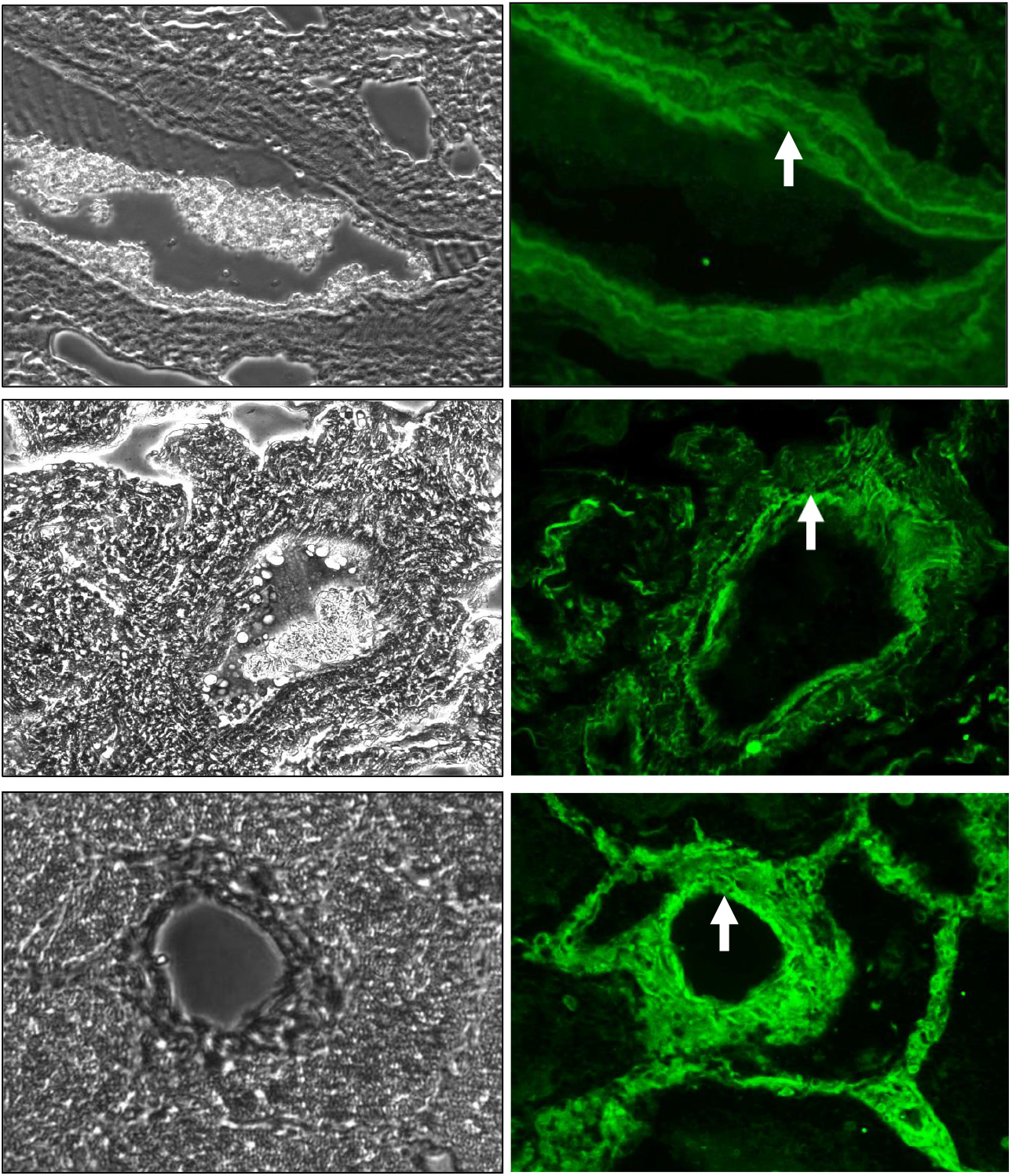
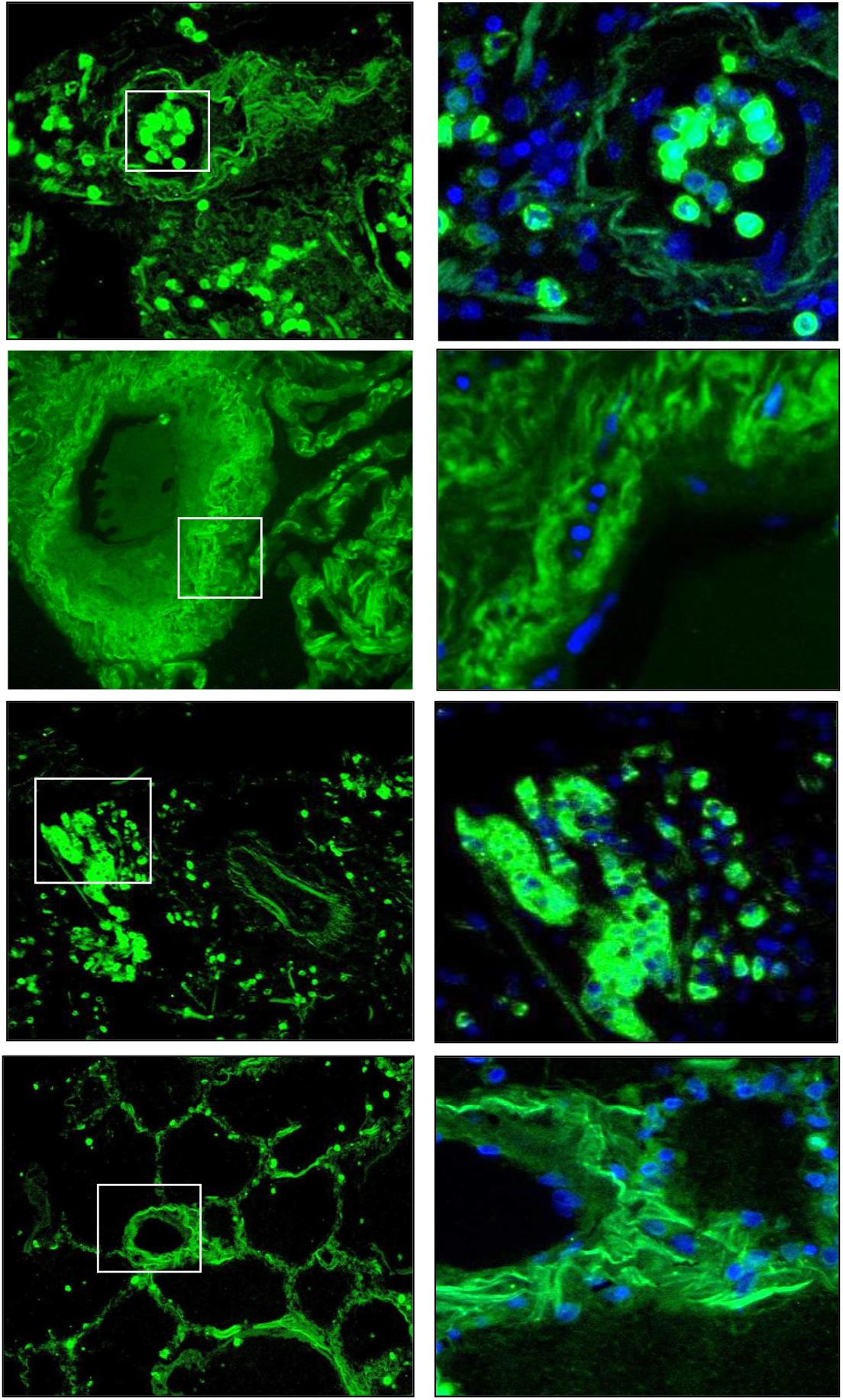
Inflammasome in the lungs of patients with COVID-19 infection. Representative images of the immunofluorescence in lung sections stained with A) anti-NLRP3 and B) caspase-1. The NLRP3 subunit (green) was visualized along the walls of arterioles (arrow). Caspase staining is widely distributed throughout the lungs.

## DISCUSSION

COVID-19 has been described largely as a respiratory disease; indeed, the respiratory tract and alveolus are amongst the primary sites of infection. However, it is also an inflammatory disease where release of inflammatory cytokines is the cause of organ injury and damage. As the endothelium is the converging site of the inflammation due to its activation (expression of adhesion molecules and cytokines) that leads to immune cell recruitment, it is reasonable to conclude that COVID-19 is potentially a vascular disease (1, 9, 21). While this would be an indirect impact of the virus, more recent studies provide evidence of a direct effect i.e. infection by SARS-CoV-2 virus of endothelial cells (24). Our inspection of autopsies of the 8 COVID-19 patients showed macro and microthrombi in almost all fields imaged, indicating coagulation pathology. As is well established, coagulation is closely linked to endothelial inflammation signaling; inflammatory moieties on the endothelium increase leukocyte infiltration and alter coagulation control driving a procoagulant direction (35). Thus, the COVID-19 which is increasingly being described as a vascular disease is perhaps more accurately defined as a pathology which has its origins in “endothelial inflammation” signaling.

Inflammasome activation on the endothelium plays a major part in cell death and injury with inflammation. The NLRP3 inflammasome is a multiprotein complex comprised of three basic components: (1) A sensor such as a NOD-like receptor (NLR) (2) the adaptor protein apoptosis-associated speck-like protein containing a caspase-recruitment domain (ASC) and (3) the inflammatory cysteine aspartase caspase-1. The assembly of this complex leads to release of an active form of caspase-1 which then exerts its catalytic activity on the pro-inflammatory cytokines (IL-1β) that after their release perpetuate cell death, specifically inflammation induced cell death or pyroptosis (3, 31).

A recent report showed high levels of NLRP3 inflammasome and caspase-1 in patients with fatal COVID (34). This is not surprising as increased NLRP3 is associated with various inflammatory lung pathologies including acute lung injury and ARDS (12, 20). The COVID-19 lung autopsies in this study, showed NLRP3 expression throughout the lung, but intense expression was seen along the lung vessel walls implying inflammasome expression on the endothelium. Caspase-1 expression was observed throughout the lungs including in the vascular structures. Caspase-1 is considered as a key pyroptotic mediator; it reportedly drives pulmonary vascular endothelial cell death (31). Elsewhere too, high caspase-1 expression has been reported with both COVID-19 (34) and with other lung inflammatory pathologies (30); however its expression on the endothelium or vascular wall with COVID-19 has not been documented. Possibly the NLRP3-caspase-1 axis can directly (via pyroptosis) or indirectly (via NLRP3 driven chemotactic immune cell recruitment (17)) injure the endothelial layer. This confluence of vascular injury, thrombosis and dysregulated inflammation seems to propagate lung damage with COVID-19 and supports a pivotal role for the pulmonary endothelium in severe and fatal COVID-19.

As NLRP3 inflammasome driven pyroptosis is being considered to play a leading role in the pathogenesis of multi-organ failure with COVID-19 (22), there is some speculation on the mechanisms by which inflammasome activation occurs upon SARS-CoV-2 infection. One possibility is that the SARS-CoV-2 spike protein’s binding to cell surface-expressed angiotensin-converting enzyme 2 (ACE2) directly triggers its enzymatic activation and alters membrane polarity that can result in activation of NLPR3 inflammasome (18). Or NLRP3 could be activated via Angiotensin II which is reported to facilitate the assembly of the inflammasome. A third possibility could be via interaction of damage associated molecular patterns (DAMPs that are released post infection) and members of the complement cascade with the SARS-CoV-2 virus. Potent cleavage fragments of DAMPs and complement cascade can potentially activate the inflammasome (26). Yet another possibility is that the stretch from ventilation activates the inflammasome (38). Once activated, the NLRP3 signaling cascade would lead to release of caspase-1 and interleukin-1β that would facilitate pyroptosis.

To the best of our knowledge, this is the first study on NLRP3 expression in the vascular structures in lungs of fatal cases of COVID-19. The origin of several events that exacerbate inflammation and injury with COVID-19 (such as immune cell aggregation and extravasation, edema, formation of thrombi and leukopenia) possibly lies in pulmonary endothelial inflammasome activation and pyroptotic cell death. Therefore, NLRP3 inhibitors have been suggested for as a potential treatment strategy and are currently being explored for management of moderate COVID-19 symptoms (NCT04540120) (11, 27).

A major drawback of this study is that our sample size is small. Moreover, paraffin based post mortem samples offer a snapshot of the disease and cannot recreate the evolving disease process. Histology is also impacted with the effects of clinical care and treatment as comorbidities, ventilation and medication pose as challenges in interpretation of results. Nevertheless, this study identifies endothelial NLRP3 inflammation, and documents thrombi and altered vascular structures in the lungs of fatal COVID-19 patients. Overall, this report adds to the growing list of studies on COVID-19 associated pulmonary pathology that highlight the importance of vascular endothelial inflammation in progression to severe and fatal disease.

## Data Availability

The raw data is available with the coresponding author and can be shared with interested parties.

